# Effectiveness of second booster compared to first booster and protection conferred by previous SARS CoV-2 infection against symptomatic Omicron BA.2 and BA.4/5 in France

**DOI:** 10.1101/2023.01.11.23284137

**Authors:** Cynthia Tamandjou, Vincent Auvigne, Justine Schaeffer, Sophie Vaux, Isabelle Parent du Châtelet

**Author notes:** These authors contributed equally to this work (Cynthia Tamandjou, Vincent Auvigne).

## Abstract

In face of evidence of rapid waning of vaccine effectiveness against Omicron and its sub-lineages, a second booster with mRNA vaccines was recommended for the most vulnerable in France. We used a test negative design to estimate the effectiveness of the second booster relative to the first booster and the protection conferred by a previous SARS-CoV-2 infection, against symptomatic Omicron BA.2 or BA.4/5. We included symptomatic ≥60 years old individuals tested for SARS-CoV-2 in March 21-October 30, 2022. Compared to a 181-210 days old first booster, a second booster restored protection with a relative effectiveness of 39% [95%CI: 38% - 41%], 7-30 days post-vaccination This gain in protection was lower than the one observed with the first booster, at equal time points since vaccination. High levels of protection were associated to previous SARS-CoV-2 infection, especially if the infection was recent and occurred when an antigenic-related variant was dominant.

**Highlights:**

- A second Wuhan-like mRNA booster brought additional protection against symptomatic Omicron BA.2 or BA.4/5 infections, relative to a first booster given 181 to 210 days ago.
- The gain in protection offered by a second booster was lower to the protection observed with a first booster, at equal time points since these booster doses.
- Previous infection, in a vaccinated population, offered high levels and long-lasting protection against symptomatic Omicron BA.2 or BA.4/5 infections.

## Introduction

Since December 2021, the Omicron (B.1.1.529) variant and its subsequent sub-lineages have driven the on-going COVID-19 pandemic. This variant has been responsible for the highest number of SARS-CoV-2 infections recorded globally to date, due to its intrinsic greater transmissibility and immune escape properties. Indeed, real-world evidence has demonstrated rapid waning of vaccine effectiveness against this variant and its sub-lineages **[1, 2];** which prompted recommendations for a fourth dose (second booster).

Despite high coverage of vaccination, the important Omicron epidemic experienced by the French population resulted in a large pool of individuals with potential immunity induced by both vaccination and infection **[3]**. In March 2022, amidst the Omicron BA.2 epidemic, the second booster dose became recommended for the most vulnerable people including 80 years and older, from three months after receiving the first booster dose. The evolving epidemic led the French authorities to extend this booster dose to people aged 60-79 years, from six months following their first booster dose. A better understanding of the effect of the second booster, amidst the current Omicron BA.4/5 epidemic is needed going forward in the pandemic. As vaccination coverage with a first booster is now high in this population, absolute vaccine effectiveness (compared to an unvaccinated population) was difficult to estimate.

### Objectives

This study examined the (1) relative effectiveness of the second Original mRNA booster dose compared with the first booster dose, and (2) protection conferred by previous SARS-CoV-2 infection against Omicron BA.2 and Omicron BA.4/5 symptomatic infection among vaccinated individuals.

## Methods

We conducted a retrospective test-negative design study on ≥60 years-old individuals with self-reported COVID-19-like symptoms, and a SARS-CoV-2 test (RT-PCR or an antigen test) performed between March 21 and October 30, 2022; period during which BA.2 and then BA.4/5 were the dominant variants in France. Eligible individuals had received at least three doses of vaccine consisting of a 2-doses primary vaccination series and one booster, and presented with no immunocompromising conditions. SARS-CoV-2 diagnostic and vaccination data were extracted from the French national information systems for all SARS-CoV-2 tests (SI-DEP), and for COVID-19 vaccine status and comorbidities (VAC-SI). Individuals with a positive test were considered as cases, and those with a negative one were controls. Cases and controls were matched in a 1:2 ratio according to the week of SARS-CoV-2 testing, type of test (antigen or RT-PCR) and area of residence (department).

Demographic characteristics as well as previous infection status data were collected. Previous SARS-CoV-2 infection was defined as a documented positive PCR or antigen test identified at least 60 days prior to the diagnostic test of inclusion, irrespective of the presence of symptoms. Variants of current infection were attributed based on (1) individual-based mutation screening performed with a multiplex RT-qPCR targeting a set of predefined mutations associated to Omicron BA.2 and Omicron BA.4/5, where available; or (2) period of circulation of a dominant variant (variant circulating at least at a 90% prevalence), determined using representative genomic surveillance data at regional level. The variant of previous infection was the dominant variant at the date of the positive test associated to the aforesaid infection. Periods during which no variant was dominant are defined as transition periods. Vaccination status was assessed at the time of the SARS-CoV-2 test. Booster 1 status was defined as complete primary vaccination (with two vaccine doses) plus one mRNA booster dose, and booster 2 status was complete primary vaccination with two mRNA booster doses.

Conditional logistic regression was used to compare the odds of testing positive for SARS-CoV-2 with one booster or two booster doses, with or without previous infection, among cases and controls. The odds ratios (OR) were estimated according to the time elapsed since (i) the last booster dose and (ii) the previous infection, and (iii) the dominant variant at the time of the previous infection. Equal time intervals since the last booster dose were used: 7-30, 31-60, 61-90, 91-120, 121-150, 151-180 and 181-210 days; allowing to measure the duration of protection induced by the second booster over time. Results associated to previous infection during period of transitions are not presented. The reference group represented individuals who received one booster dose and had 181-210 days passed since this vaccine dose (booster 1). The OR were adjusted on sex, age group (50-79 and ≥80 years old), residence type (individual housing or Long Term Care Facilities [LTCFs]), presence of medium-risk comorbidities (yes or no), and total number of SARS-CoV-2 tests in the last six months (used as a proxy of healthcare seeking behaviour). The relative vaccine effectiveness (rVE) of the second booster compared to the first booster, and protection conferred by previous SARS-CoV-2 infection **[4]** were derived as [1-adjusted OR (aOR)] x 100 when OR≤1 and as [1/OR – 1] x 100 when OR > 1 with 95% CI. Estimations were made for each time interval since vaccination. Statistical analyses were performed using R, version 4.1.3. Further details are provided in the supplementary appendix.

## Results

In total, 933 491 individuals were included in the analysis; of whom 456 657 (49%) SARS-CoV-2 positive (cases) were matched to 476 834 (51%) SARS-CoV-2 negative (controls) (Supplementary appendix, Figure S2). Majority of participants were in the 60-79 age group (80%), and had received only one booster dose (72%, Table 1). Moreover, 92% had no documented previous SARS-CoV-2 infection. Fifty-one percent (51%) of positive SARS-CoV-2 tests were attributed to Omicron BA.2, 36% occurred during the period of transition from BA.2 to BA.4/5, and 13% were allocated to Omicron BA.4/5 (Figure S3). The majority of individuals (74%) had received the mRNA BNT162b2 BioNTech/Pfizer vaccine as booster 1, compared to 26% for mRNA-1273 Moderna vaccine. Similarly, among those with a second booster, 85% were vaccinated with the BioNTech/Pfizer vaccine and 15% with the Moderna vaccine (Table 2).

**Table 1:**
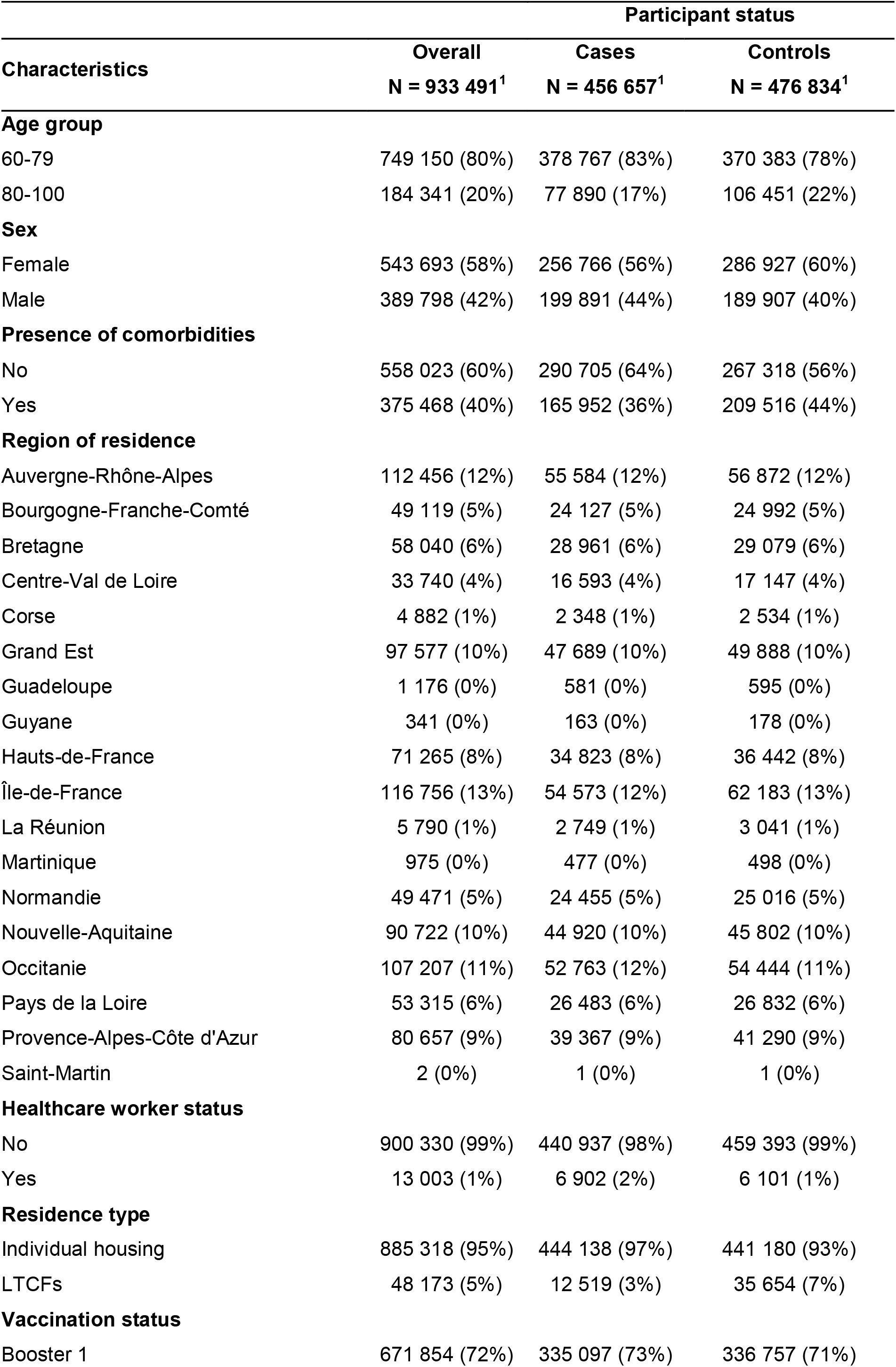
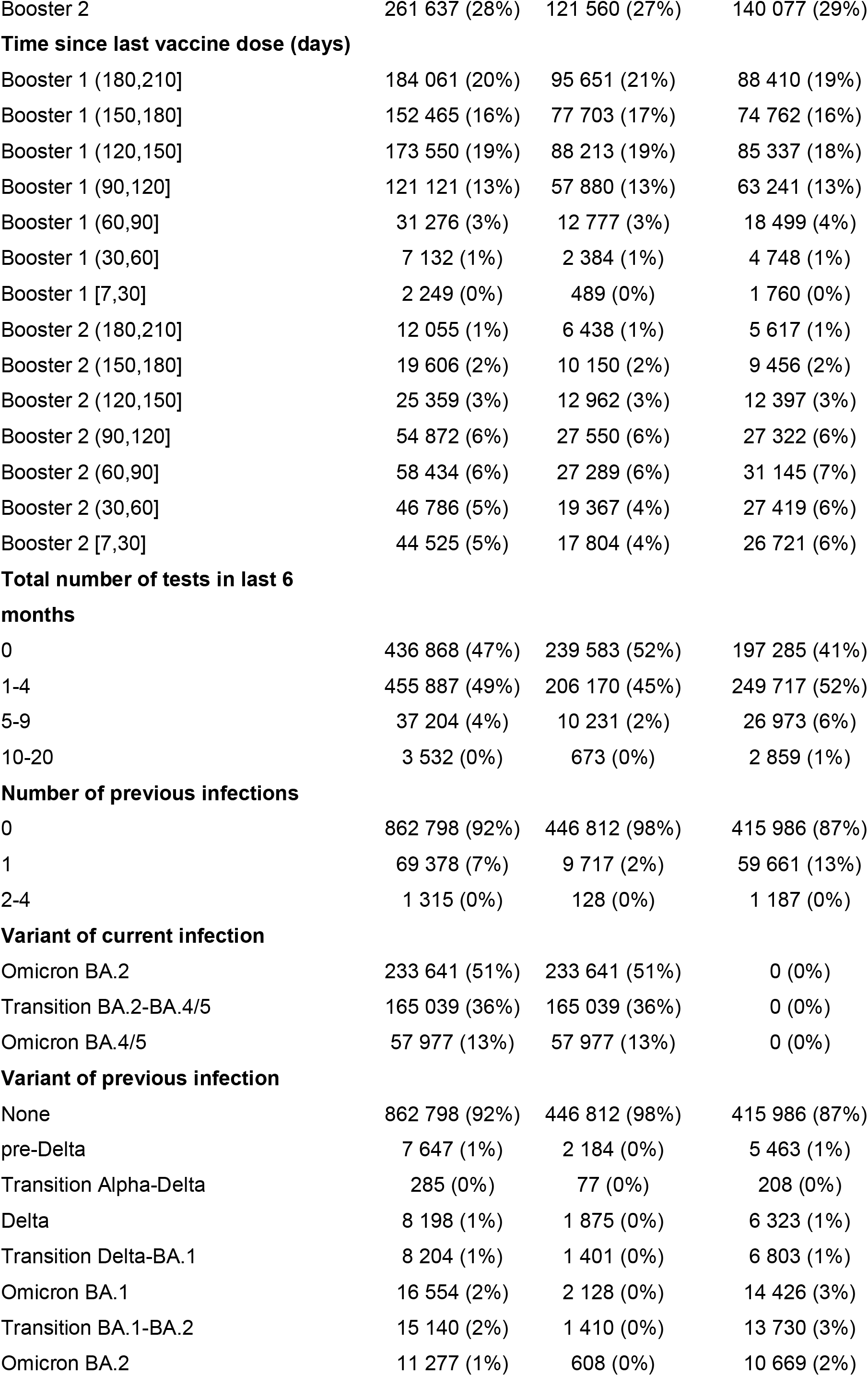
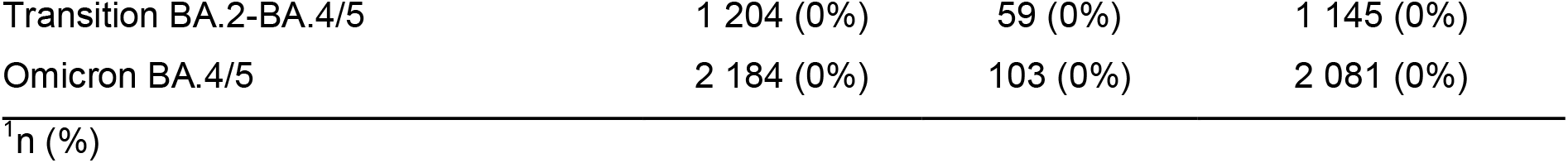
Descriptive statistics of the study population.

**Table 2:** Distribution of vaccine types according to the vaccination status.

| Vaccine types | Vaccination status |  |
| --- | --- | --- |
|  | Booster 1<br>N = 671 854<sup>†</sup> | Booster 2<br>N = 261 637<sup>†</sup> |
| **Primary 1** |  |  |
| Pfizer Original | 463 914 (69%) | 186 561 (71%) |
| AstraZeneca | 145 456 (22%) | 54 870 (21%) |
| Moderna Original | 62 482 (9%) | 20 206 (8%) |
| Novavax | 2 (0%) | 0 (0%) |
| **Primary 2** |  |  |
| Pfizer Original | 470 510 (70%) | 188 943 (72%) |
| AstraZeneca | 135 306 (20%) | 51 648 (20%) |
| Moderna Original | 66 032 (10%) | 21 046 (8%) |
| Novavax | 6 (0%) | 0 (0%) |
| **Booster 1** |  |  |
| Pfizer Original | 499 539 (74%) | 222 059 (85%) |
| Moderna Original | 172 277 (26%) | 39 562 (15%) |
| AstraZeneca | 28 (0%) | 16 (0%) |
| Novavax | 10 (0%) | 0 (0%) |
| **Booster 2** |  |  |
| Pfizer Original | 0 (0%) | 248 410 (95%) |
| Moderna Original | 0 (0%) | 13 194 (5%) |
| Novavax | 0 (0%) | 33 (0%) |
| <sup>†</sup>n (%) |  |  |

Original = the monovalent Wuhan-like vaccine.

Estimates of the adjusted rVE and aOR by days since administration of the first and second boosters are presented in Figure 1 and Table S1 (Supplementary appendix), respectively. Compared with individuals who received the first booster dose 181-210 days before being tested, the relative protection associated to the first booster and the second booster 7-30 days post vaccination was 64% [95% CI: 60% - 68%] and 39% [38% - 41%], respectively. A statistically significant decrease in the rVE, across time intervals since last vaccination, was observed for both booster 1 and booster 2. However, at similar vaccination time points, the gain in protection measured by the rVE offered by the second booster was lower than the protection offered by the first booster. For instance, rVE of the second booster 91-120 days ago was 8% [5% - 10%] whereas at the same time interval, the rVE of the first booster was 33% [32% - 35%] (Figure 1).

**Figure 1:**
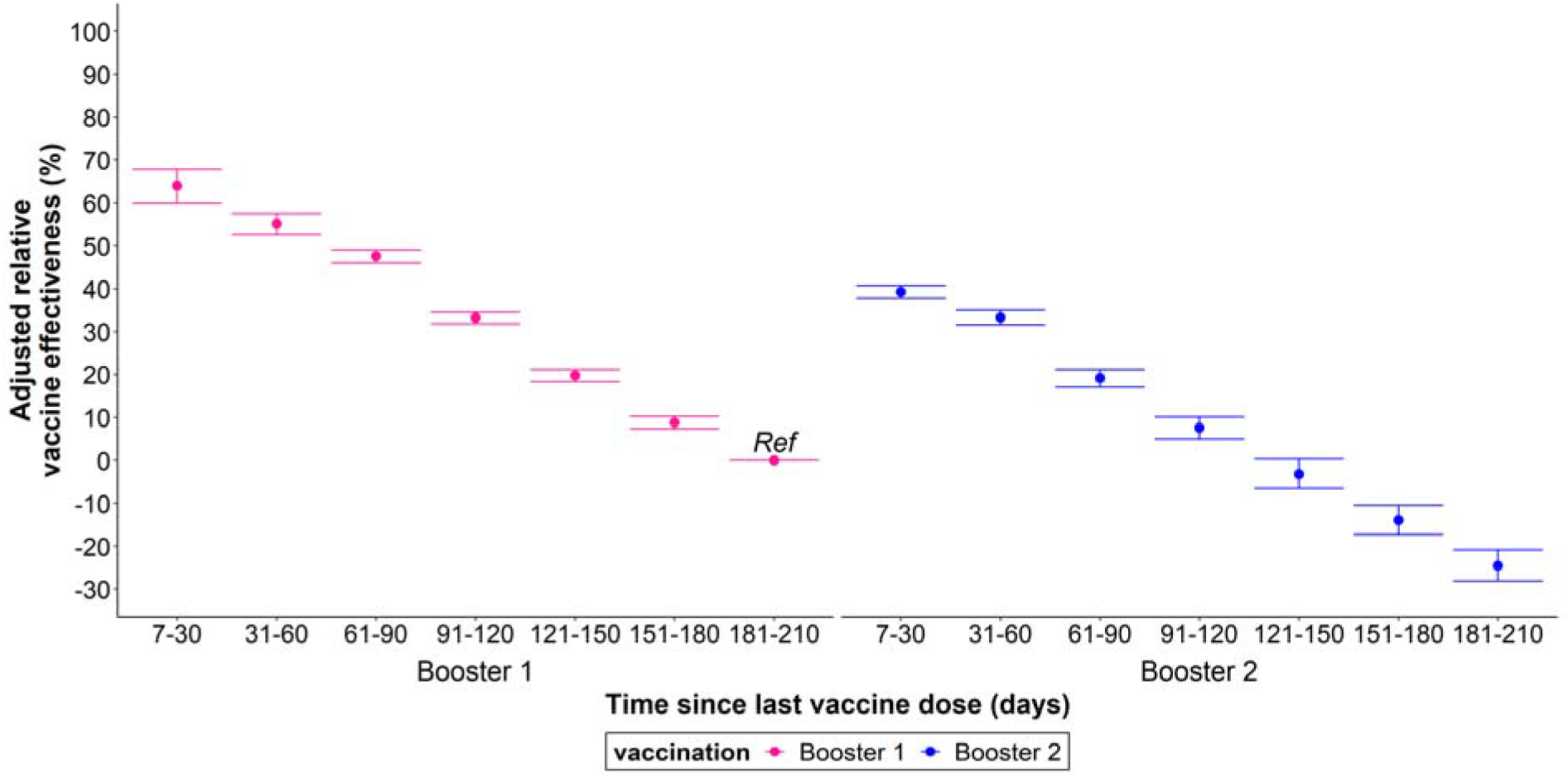
Adjusted relative vaccine effectiveness of the second booster of mRNA COVID-19 vaccine, compared to those who received the first booster dose 181-210 days previously, against symptomatic Omicron BA.2 and BA.4/5 infection among ≥60 years and older. Error bars = 95% confidence intervals of the estimates.

Compared to people without previous infection, the more recent was the previous infection, the higher was the protection against symptomatic infection (Figure 2). For instance, the adjusted protection associated with a 61-112 days old previous infection which occurred during the Omicron BA.2 dominant period was 95.6% [95.0%–96.1%] whereas a 321-467 days old previous infection from the Delta-predominant period was associated with protection of 61.7% [57.5%–65.5%]. Note that comparing the protection induced by the different dominant variants of previous infections can only be done when data are available at similar time intervals since previous infection. For instance, five months (≥150 days) following a previous infection, protection was higher if the previous infection occurred during the BA.2 dominant period compared to during the BA.1 period (Figure 2, Table S1): 90.9% [89.5%–92.2%] vs 78.6% [76.5%–80.4%], respectively.

**Figure 2:**
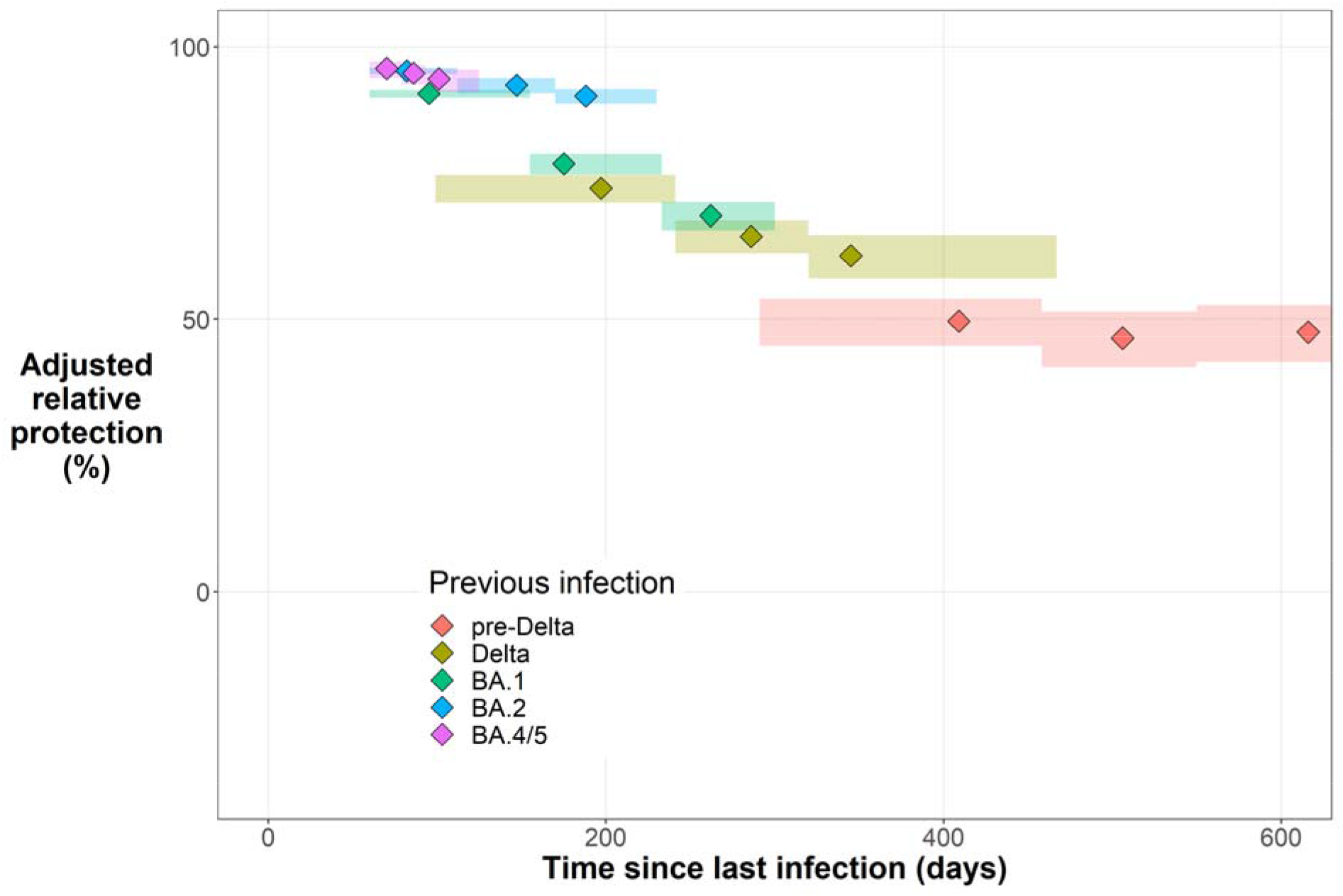
Adjusted protection associated with a previous SARS-CoV-2 infection against Omicron BA.2 and BA.4/5 symptomatic infection among ≥60 years old. Protection is estimated according to the time since and variant dominant at previous infection. Individuals without a documented previous infection served as reference. Rectangles around the point estimates represent the time range (horizontally), and the 95% CI of the estimates (vertically). Periods of transitions between variants are not represented.

## Discussion

To date, few studies have explored the protection gained from the second COVID-19 booster vaccines. In this test-negative case-control study, we found that a recent second booster dose of BNT162b2 or mRNA-1273 vaccine provided additional protection against symptomatic COVID-19 infection, relative to the residual protection of a 6-month-old (180-210 days) first booster dose. This was observed when Omicron BA.2 and BA.4/5 were dominant in France. Similar observations have been reported in recent studies from Israel **[5, 6]** and Canada **[7]**. In Israel, the rVE against infection of a second booster, compared to the first one received at least four months ago, among adults aged 60 years and older was estimated at 58% [56% - 60%] 7-13 days from vaccination. This VE quickly declined over time to 22% [5% - 36%] at 63-69 days **[6]**. The Canadian study also found that, compared to the first booster dose of mRNA vaccine received ≥84 days ago, the second booster offered additional protection against any SARS-CoV-2 infection and symptomatic infection, estimated at 19% [12% - 26%] and 31% [20% - 41%], respectively among long-term care residents aged ≥60 years **[7]**. It is important to mention that this gain in protection provided by the second booster adds to a residual protection provided by the first booster. Although we could not measure the absolute VE of an old first booster compared to unvaccinated individuals, this estimate has been reported elsewhere. Link-Gelles and colleagues in the USA estimated this VE at 32% [26% - 38%] against laboratory-confirmed COVID-19–associated emergency department or urgent care encounters among adults ≥50 years old, ≥120 days post receiving the first booster (third vaccine dose), during the Omicron BA.2–predominant period **[8]**.

Although additional protection with the second booster dose was achieved, it decreased over time reaching levels below 10% at >90 days post vaccine administration. Indeed, waning of protection against SARS-CoV-2 infection is consistent with observations made with previous COVID-19 vaccine doses, especially amidst the Omicron epidemic **[2, 9]**. Nonetheless, we noted that, compared to a 6-7 month old first booster, the gain in protection offered by the second booster was significant but lower than the protection offered by the first booster, at similar vaccination time points. Supplementary analyses showed similar patterns for both Omicron BA.2 and BA.4/5 periods of inclusion (Figure S4), and between age groups (Figure S5). This moderate gain in protection may be explained by a high baseline protection conferred by the first booster dose. This hypothesis goes along data from an open-label, nonrandomized clinical study conducted in Israel among healthcare workers. The participants had received four doses of either BNT162b2 Pfizer– BioNTech or mRNA-1273 Moderna vaccines four months after the third dose in a series of three BNT162b2 doses. Results from the study suggested that three doses achieves maximal immunogenicity of mRNA vaccines; titers of vaccines induced IgG antibodies and neutralizing antibody titers were slightly higher after receiving the fourth dose, than those achieved after the third dose **[10]**. We may also bring forward the hypothesis of immune imprinting **[11]**. So far, the vaccinated population has been repeatedly stimulated with vaccines targeting the conserved regions of the Original spike protein, therefore enabling a strong and specific immune response against this virus and other closely related antigenic variants. In the context of immune imprinting, frequent exposure to the same antigen is to the detriment of new neutralizing responses against variant antigens. The Omicron variant differs substantially from this wild-type virus, with a greater number of mutations within the receptor-binding region **[12]**. A few reports have eluded to this phenomenon in the context of vaccine effectiveness against Omicron infection, in presence and in absence of previous infection **[13-15]**. Moreover, it has been shown in animal model that repeated boosters induce humoral and cellular immune tolerance **[16]**. Still, there is a need for human clinical studies to evaluate the impact of immune imprinting, amidst this evolving pandemic and the advent of new SARS-CoV-2 variants. These data would be important to inform future vaccination policies, and to understand the effects of new bivalent vaccines.

This study also showed that having experienced a recent previous infection was associated with high levels of protection against symptomatic infection, more so during the period of dominance of an antigenic-related variant. Similar observations have been previously reported in France **[3]**, and elsewhere **[4]**. Interestingly, protection remained high even among individuals who experienced a non-Omicron previous infection. Protection associated with a 321-467 days old previous infection from the Delta-predominant period was 61.7% [57.5%–65.5%]. Almost two years after the previous infection, moderate protection remained among these vaccinated individuals. This observation reflects the strength of the combined effects of infection and vaccination also known as hybrid immunity **[17, 18]**.

This analysis presents limitations to consider. Firstly, unrecognized or undocumented symptomatic COVID-19 positive infection may have influenced vaccine-induced protection in this population. The availability of SARS-CoV-2 self-tests which are not captured in SI-DEP may have reduced the detection of COVID-19 positive tests. Secondly, despite the matching and controlling for various covariates, some residual or unmeasured confounding may remain. For example, being vaccinated may influence the decision to perform a SARS-CoV-2 test at onset of symptoms related to the disease. Similarly, the severity of symptoms may also influence the decision of testing. Individuals with light symptoms, especially in the few days following vaccination, may not get tested whereas those with severe symptoms may do so. To limit this bias, individuals within similar time points since vaccination were compared. Moreover, the lack of data related to socio-economic status did not allow the consideration of this potential confounder, which may influence the decision to be vaccinated and the access to vaccines. Thirdly, data on previous infections do not include undiagnosed previous infections, which may have led to an underestimation of the marginal protection conferred by previous infection.

## Conclusion

Findings from this study show additional protection with a second booster dose of Original mRNA vaccines, compared to a 6-7 month old first booster dose. However, the gain in protection offered by a second booster was inferior to the protection observed with a first booster at equal time since these booster doses. Moreover, in the context of an Omicron-driven pandemic, vaccination coupled with recent prior infection offer high levels of protection against subsequent Omicron infection. It will be interesting to understand the contribution of bivalent vaccines in this evolving pandemic.

## Data Availability

While all data used in this analysis were pseudonymised, the individual-level nature of the data used risks individuals being identified, or being able to self-identify, if it is released publicly. Requests for access to the underlying source data should be directed to Sante publique France and will be granted in accordance with the GDPR and French Law.

## Ethical statement

This study did not involve the human person. It was carried out by Santé Publique France using data collected by the Ministry of Health to manage the COVID-19 crisis. This processing of personal pseudonymised data was implemented in accordance with the legislative and regulatory prerogatives granted to Santé publique France to fulfil its public interest mission and in compliance with the provisions of the GDPR. In this context, the opinion of an ethics committee was not required.

## Authors contributions

CT, VA, JS, SV and IPC conceived and designed the study; CT and VA□reviewed the literature, □managed the datasets, did the statistical analysis and wrote the original draft. All authors contributed to the discussion, reviewed the manuscript and made the decision to submit for publication.

## Acknowledgments

We would like to thank the health care professionals and administrative teams in hospitals and biomedical laboratories, the professionals in charge of vaccinations and all those who contributed to the data management and analysis over the past three years. We would particularly like to thank Bruno Coignard and Didier Che, whose scientific and organisational support made this study possible, Daniel Levy-Bruhl, who set up the vaccine effectiveness studies in our team, Yann Le Strat, for his support in biostatistics and Charline Montagnat (Sully group) for data processing.

## Data sharing statement

Although the data used in this analysis has all been pseudonymised, there remains a risk that individuals could be identified, or could self-identify, if released publicly, due to the individual nature of the data used. Requests for access to the underlying source data should be directed to Santé publique France and will be granted in accordance with the GDPR and French Law.

## References

[1] Andrews N, Stowe J, Kirsebom F, Toffa S, Rickeard T, Gallagher E, et al. Covid-19 Vaccine Effectiveness against the Omicron (B.1.1.529) Variant. N Engl J Med. 2022;386:1532–46. https://doi.org/10.1056/NEJMoa2119451

[2] Castillo MS, Khaoua H, Courtejoie N. Vaccine effectiveness and duration of protection against symptomatic infections and severe Covid-19 outcomes in adults aged 50 years and over, France, January to mid-December 2021. Global Epidemiology. 2022:100076. https://doi.org/10.1016/j.gloepi.2022.100076

[3] Auvigne V, Schaeffer J, Boudon T, Tamandjou C, Figoni J, du Châtelet IP, et al. Risk of SARS-CoV-2 reinfection is time- and variant-dependant, France, January 2021 to August 2022. medRxiv. 2022:2022.11.09.22282113. https://doi.org/10.1101/2022.11.09.22282113

[4] Carazo S, Skowronski DM, Brisson M, Barkati S, Sauvageau C, Brousseau N, et al. Protection against Omicron BA.2 reinfection conferred by primary Omicron or pre-Omicron infection with and without mRNA vaccination. medRxiv. 2022:2022.06.23.22276824. https://doi.org/10.1101/2022.06.23.22276824

[5] Magen O, Waxman JG, Makov-Assif M, Vered R, Dicker D, Hernán MA, et al. Fourth Dose of BNT162b2 mRNA Covid-19 Vaccine in a Nationwide Setting. New England Journal of Medicine. 2022;386:1603–14. https://doi.org/10.1056/NEJMoa2201688

[6] Gazit S, Saciuk Y, Perez G, Peretz A, Pitzer VE, Patalon T. Short term, relative effectiveness of four doses versus three doses of BNT162b2 vaccine in people aged 60 years and older in Israel: retrospective, test negative, case-control study. BMJ. 2022;377:e071113. https://doi.org/10.1136/bmj-2022-071113

[7] Grewal R, Kitchen SA, Nguyen L, Buchan SA, Wilson SE, Costa AP, et al. Effectiveness of a fourth dose of covid-19 mRNA vaccine against the omicron variant among long term care residents in Ontario, Canada: test negative design study. BMJ. 2022;378:e071502. https://doi.org/10.1136/bmj-2022-071502

[8] Link-Gelles R, Levy ME, Gaglani M, Irving SA, Stockwell M, Dascomb K, et al. Effectiveness of 2, 3, and 4 COVID-19 mRNA Vaccine Doses Among Immunocompetent Adults During Periods when SARS-CoV-2 Omicron BA.1 and BA.2/BA.2.12.1 Sublineages Predominated - VISION Network, 10 States, December 2021-June 2022. MMWR Morb Mortal Wkly Rep. 2022;71:931–9. https://doi.org/10.15585/mmwr.mm7129e1

[9] Tartof SY, Slezak JM, Fischer H, Hong V, Ackerson BK, Ranasinghe ON, et al. Effectiveness of mRNA BNT162b2 COVID-19 vaccine up to 6 months in a large integrated health system in the USA: a retrospective cohort study. Lancet. 2021;398:1407–16. https://doi.org/10.1016/S0140-6736(21)02183-8

[10] Regev-Yochay G, Gonen T, Gilboa M, Mandelboim M, Indenbaum V, Amit S, et al. Efficacy of a Fourth Dose of Covid-19 mRNA Vaccine against Omicron. N Engl J Med. 2022;386:1377–80. https://doi.org/10.1056/NEJMc2202542

[11] Davenport FM, Hennessy AV, Fabisch WtTAoPH. Predetermination by infection and by vaccination of antibody response to influenza virus vaccines. The Journal of experimental medicine. 1957;106:835–50.

[12] Xu C, Wang Y, Liu C, Zhang C, Han W, Hong X, et al. Conformational dynamics of SARS-CoV-2 trimeric spike glycoprotein in complex with receptor ACE2 revealed by cryo-EM. Sci Adv. 2021;7:eabe5575. https://doi.org/10.1126/sciadv.abe5575

[13] Chemaitelly H, Ayoub HH, Tang P, Hasan MR, Coyle P, Yassine HM, et al. Immune Imprinting and Protection against Repeat Reinfection with SARS-CoV-2. New England Journal of Medicine. 2022;387:1716–8. https://doi.org/10.1056/NEJMc2211055

[14] Ju B, Fan Q, Wang M, Liao X, Guo H, Wang H, et al. Antigenic sin of wild-type SARS-CoV-2 vaccine shapes poor cross-neutralization of BA.4/5/2.75 subvariants in BA.2 breakthrough infections. Nat Commun. 2022;13:7120. https://doi.org/10.1038/s41467-022-34400-8

[15] Reynolds CJ, Pade C, Gibbons JM, Otter AD, Lin KM, Munoz Sandoval D, et al. Immune boosting by B.1.1.529 (Omicron) depends on previous SARS-CoV-2 exposure. Science. 2022;377:eabq1841. https://doi.org/10.1126/science.abq1841

[16] Gao FX, Wu RX, Shen MY, Huang JJ, Li TT, Hu C, et al. Extended SARS-CoV-2 RBD booster vaccination induces humoral and cellular immune tolerance in mice. iScience. 2022;25:105479. https://doi.org/10.1016/j.isci.2022.105479 https://doi.org/10.1056/NEJMoa2208343

[17] Carazo S, Skowronski DM, Brisson M, Sauvageau C, Brousseau N, Gilca R, et al. Estimated Protection of Prior SARS-CoV-2 Infection Against Reinfection With the Omicron Variant Among Messenger RNA-Vaccinated and Nonvaccinated Individuals in Quebec, Canada. JAMA Netw Open. 2022;5:e2236670. https://doi.org/10.1001/jamanetworkopen.2022.36670

[18] Hall V, Foulkes S, Insalata F, Kirwan P, Saei A, Atti A, et al. Protection against SARS-CoV-2 after Covid-19 Vaccination and Previous Infection. N Engl J Med. 2022;386:1207–20. https://doi.org/10.1056/NEJMoa2118691

