## Appendix for "Effectiveness of second booster compared to first booster and protection conferred by previous SARS CoV-2 infection against symptomatic Omicron BA.2 and BA.4/5 in France"

### Section S1: Methods

#### Study design and period

We conducted a retrospective test-negative design study to estimate the relative vaccine effectiveness provided by a second booster dose with Wuhan-like mRNA vaccines against Omicron BA.2 and BA.4/5 symptomatic infections. Eligible individuals were selected between March 21 and October 30, 2022 (epidemiological week 12–43); period during which Omicron BA.2, then Omicron BA.4/5 predominantly circulated in France. Individuals who reported COVID-19 like symptoms and had a positive SARS-CoV-2 test (RT-PCR or antigenic test) during the study period were compared to individuals who also reported COVID-19 like symptoms but with a negative SARS-CoV-2 test (RT-PCR or antigenic test).

#### Data sources

SARS-CoV-2 diagnostic and vaccination data were extracted from the merge of two national pseudonymized information systems. These systems were set up in the context of the COVID-19 pandemic. These include SI-DEP with all SARS-CoV-2 tests (RT-PCR and antigen tests, excluding self-tests) performed in France; and VAC-SI with COVID-19 vaccination information of the French population (vaccination status, the type of vaccines administered, vaccine brand name, number of doses administered, dates of the vaccine doses administered, and vaccine priority status).

#### Study population, and inclusion criteria

The study considered individuals 60 years and older with a SARS-CoV-2 test performed during the study period. These individuals were selected based on a set of criteria including: (i) presence of self-reported COVID-19-like symptoms, (ii) absence of highly immunosuppressive comorbidities, (iii) complete primary vaccination with two vaccine doses, and (iv) vaccination with at least one booster dose administered at least seven days and at most 210 days from the inclusion date. Eligible individuals were classified as either cases or controls based on results of SARS-CoV-2 tests. Adults with at least one (1) positive RT-PCR done within seven days of symptom's onset or ten days from a positive antigenic test performed within the seven days of symptom's onset, or (2) positive antigenic test performed within seven days of symptom's onset without a subsequent positive RT-PCR test were classified as cases. Controls were symptomatic individuals with a negative RT-PCR or antigenic test done within seven days of symptom's onset; this negative test was not preceded by a positive test performed in the same week. Where multiple negative tests were recorded within the same week for an individual, only the first test was considered. Cases could become controls and vice versa provided that the subsequent negative (for the case) or positive test (for the control) were recorded during the study period but not within the same week. Cases and controls were matched in a 1:2 ratio according to the week of testing (accounting for exposure to the similar virus), type of SARS-CoV-2 diagnostic test (antigen or RT-PCR) and area of residence (department).

**Ethical considerations**

We used pseudononymised data collected from state-mandated surveillance systems, under the legal mandate of Santé publique France who serves as the public health agency in France. The process of data pseudomysation in place is done according to the French laws and in compliance with the GDPR. This process does not permit the identification of specific individuals without additional data.

**Definition of vaccination status**

Vaccination was defined at the time of the SARS-CoV-2 test. Therefore, the vaccination status was set as receipt of vaccine least seven days before SARS-CoV-2 testing. The French booster campaign involved the mRNA vaccines Pfizer/BioNTech BNT162b2 and Moderna mRNA-1273. Thus, two vaccination statuses were considered: (1) complete primary vaccination (with two vaccine doses) plus any mRNA booster dose henceforth referred as Booster 1, and (2) complete primary vaccination with two booster doses henceforth referred as Booster 2. Vaccination status was further categorized according to the time since the last dose received. Equal time intervals were used for both booster doses: 7-30, 31-60, 61-90, 91-120, 121-150, 151-180 and 181-210 days.

**Statistical analysis**

Conditional logistic regression models, which account for the matching, were fitted to the data. The SARS-CoV-2 test result was included as the dependent variable, and vaccination status was as an independent variable. Potential confounders such as sex, age group (50-79 and ≥80 years old), residence type, presence of comorbidities (yes or no), total number of SARS-CoV-2 tests in the last six months, and previous SARS-CoV-2 infection were controlled for in the models. The presence of previous SARS-CoV-2 infection was categorized according to (1) the time since the infection, and (2) the dominant variant at the time of that infection (Figure S1). The time since previous infection was estimated from the date of the inclusion SARS-CoV-2 test. A composite variable taking into consideration these two elements (time and the dominant variant) was generated for the analyses.

Vaccine effectiveness of the second booster, compared to the first one, was the preventable fraction in the unexposed when aOR≤1 (1 – aOR) and the attributable fraction in the exposed when aOR > 1 (1/aOR – 1) [1].

Figure S1: Weekly distribution of variants of previous infection.

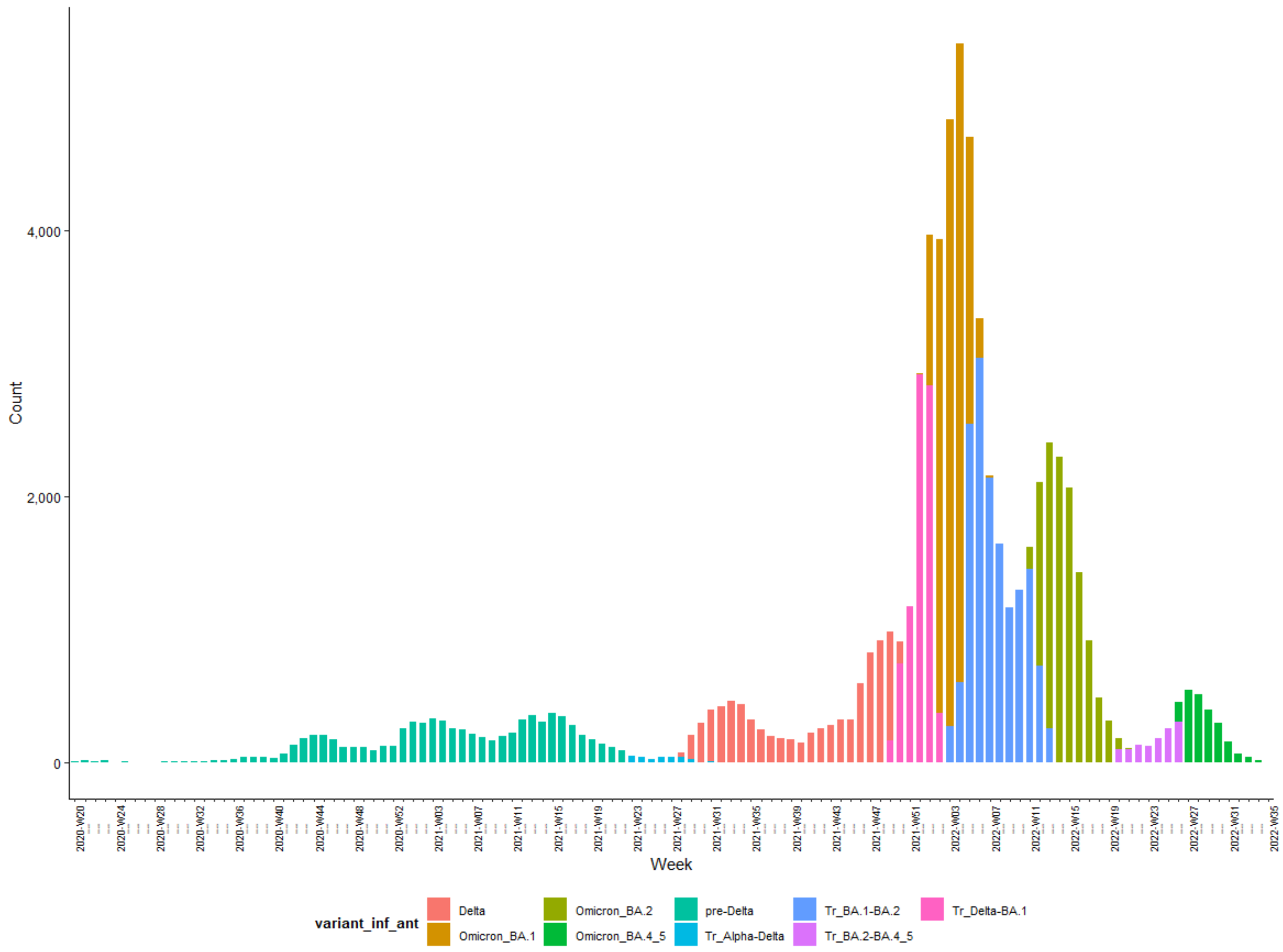

### Section S2: Results

Figure S2: Process of population selection.

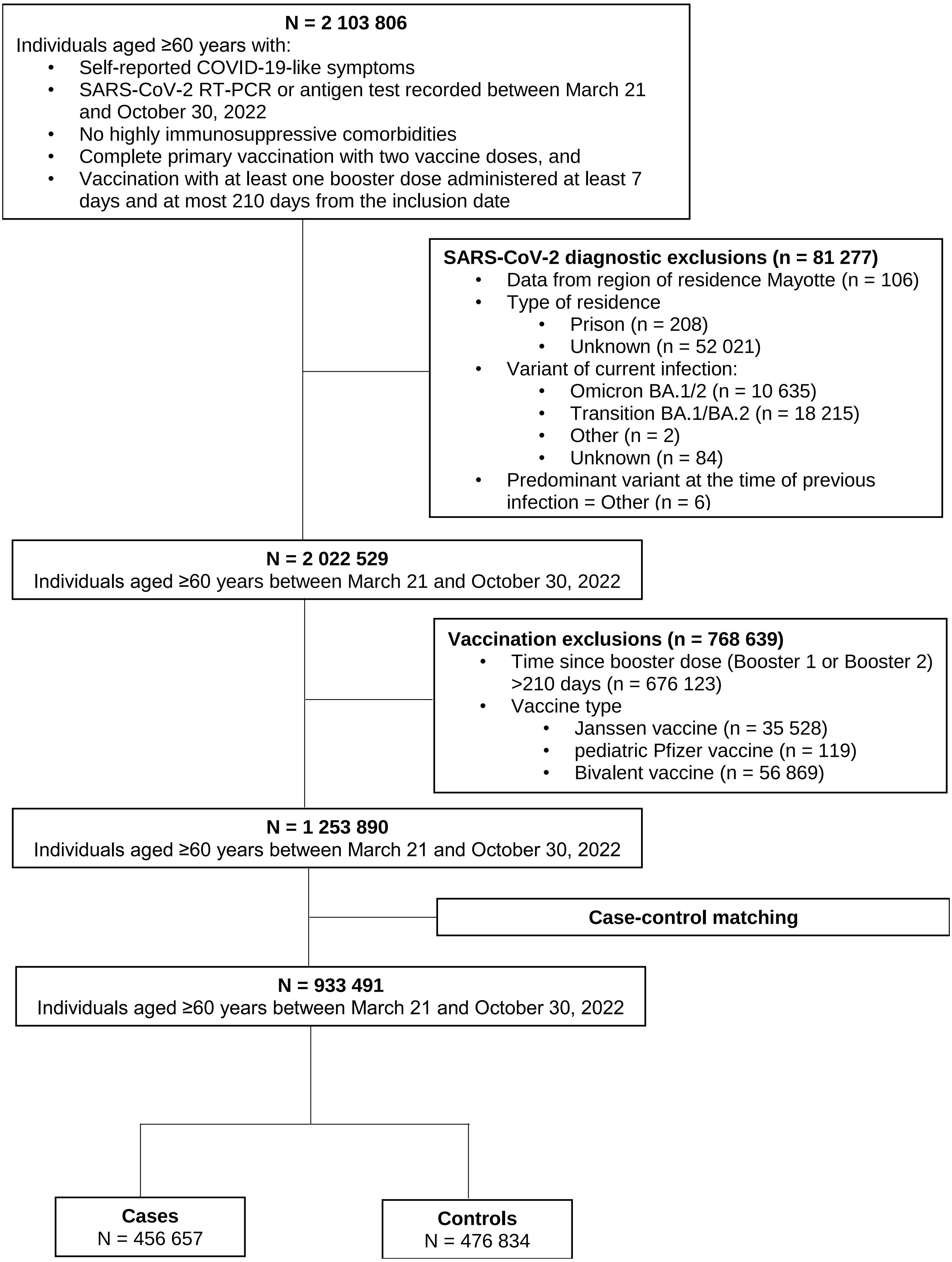

Figure S3: Number of inclusions of cases and distribution of variants across the study period.

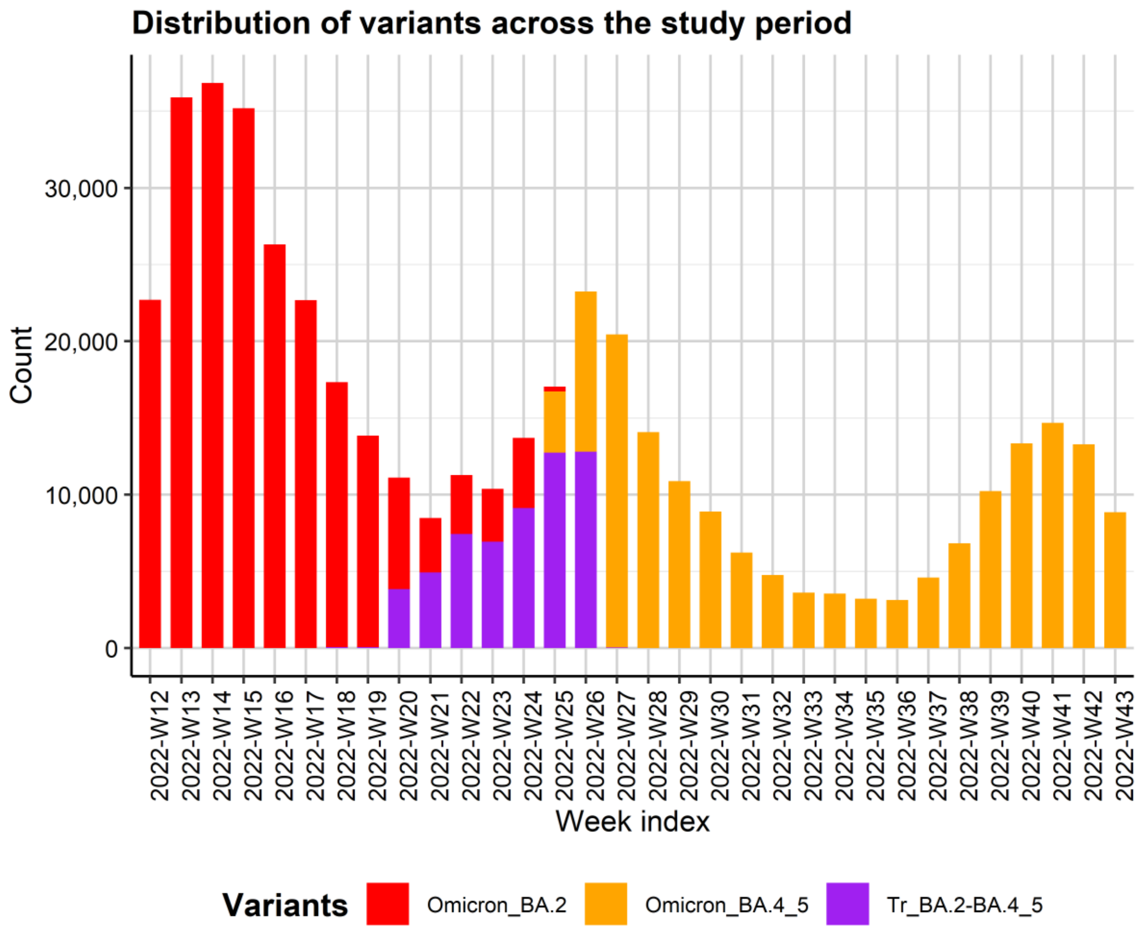


Table S1: Adjusted odds ratio and relative vaccine effectiveness (all mRNA vaccines combined) against symptomatic SARS-CoV-2 infection among ≥60 years and older.

| Variables | aOR (95% CI) | Protection against symptomatic infection (95% CI) |
|---|---|---|
| **Time since last booster dose** | | |
| Booster 1 [7,30] | 0.36 (0.32 – 0.40) | 64.04% (59.87% – 67.77%) |
| Booster 1 (30,60] | 0.45 (0.42 – 0.47) | 55.13% (52.60% – 57.53%) |
| Booster 1 (60,90] | 0.52 (0.51 – 0.54) | 47.62% (46.09% – 49.11%) |
| Booster 1 (90,120] | 0.67 (0.65 – 0.68) | 33.22% (31.86% – 34.55%) |
| Booster 1 (120,150] | 0.80 (0.79 – 0.82) | 19.73% (18.29% - 21.15%) |
| Booster 1 (150,180] | 0.91 (0.90 – 0.93) | 8.86% (7.38% – 10.31%) |
| Booster 1 (180,210] | Ref | - |
| Booster 2 [7,30] | 0.61 (0.59 – 0.62) | 39.29% (37.81% – 40.73%) |
| Booster 2 (30,60] | 0.67 (0.65 – 0.68) | 33.34% (31.65% – 35.00% |
| Booster 2 (60,90] | 0.81 (0.79 – 0.83) | 19.13% (17.03% – 21.19%) |

| | | |
|---|---|---|
| Booster 2 (90,120] | 0.92 (0.90 – 0.95) | 7.61% (4.99% – 10.16%) |
| Booster 2 (120,150] | 1.03 (1.00 – 1.07) | -3.16% (-6.51% – 0.31%) |
| Booster 2 (150,180] | 1.16 (1.12 – 1.21) | -13.92% (-17.29% – -10.42%) |
| Booster 2 (180,210] | 1.33 (1.26 – 1.39) | -_24.59% (-_28.14% – -20.86%) |
| **Age group** | | |
| 60-79 | Ref | - |
| 80-100 | 0.78 (0.77 – 0.79) | |
| **Residence type** | | |
| Individual housing | Ref | - |
| LTCFs | 0.45 (0.44 – 0.46) | |
| **Sex** | | |
| Female | Ref | - |
| Male | 1.23 (1.22 – 1.24) | |
| **Variant and time since previous infection** | | |
| None | Ref | - |
| pre-Delta (550,890] | 0.52 (0.47 – 0.58) | 47.64% (42.18% – 52.59%) |
| pre-Delta (458,550] | 0.53 (0.49 – 0.59) | 46.55% (41.25% – 51.38%) |
| pre-Delta [291,458] | 0.50 (0.46 – 0.55) | 49.63% (45.15% – 53.74%) |
| Delta (320,467] | 0.38 (0.35 – 0.43) | 61.69% (57.48% – 65.48%) |
| Delta (241,320] | 0.35 (0.32 – 0.38) | 65.23% (62.06% – 68.13%) |
| Delta [99,241] | 0.26 (0.24 – 0.29) | 74.04% (71.33% – 76.50%) |
| Omicron_BA.1 (233,300] | 0.31 (0.29 – 0.34) | 69.00% (66.29% – 71.49%) |
| Omicron_BA.1 (155,233] | 0.21 (0.20 – 0.23) | 78.55% (76.54% – 80.39%) |
| Omicron_BA.1 [60,155] | 0.09 (0.08 – 0.09) | 91.41% (90.65% – 92.11%) |
| Omicron_BA.2 (170,230] | 0.09 (0.08 – 0.11) | 90.94% (89.49% – 92.20%) |
| Omicron_BA.2 (112,170] | 0.07 (0.06 – 0.09) | 92.95% (91.44% – 94.20%) |
| Omicron_BA.2 [60,112] | 0.04 (0.04 – 0.05) | 95.59% (94.99% – 96.11%) |
| Omicron_BA.4/5 (93,125] | 0.06 (0.04 – 0.08) | 94.12% (91.67% – 95.85%) |
| Omicron_BA.4/5 (79,93] | 0.05 (0.03 – 0.07) | 95.16% (93.12% – 96.60%) |
| Omicron_BA.4/5 [60,79] | 0.04 0.03 – 0.06) | 96.03% (94.25% – 97.26%) |
| **Total number of tests in last 6 months** | | |
| 0 | Ref | - |
| 1-4 | 0.78 (0.78 – 0.79) | |
| 5-9 | 0.49 (0.48 – 0.51) | |
| 10-20 | 0.35 (0.32 – 0.38) | |

| | | |
|---|---|---|
| **Presence of comorbidities** | | |
| No | Ref | - |
| Yes | 0.74 (0.74 – 0.75) | |

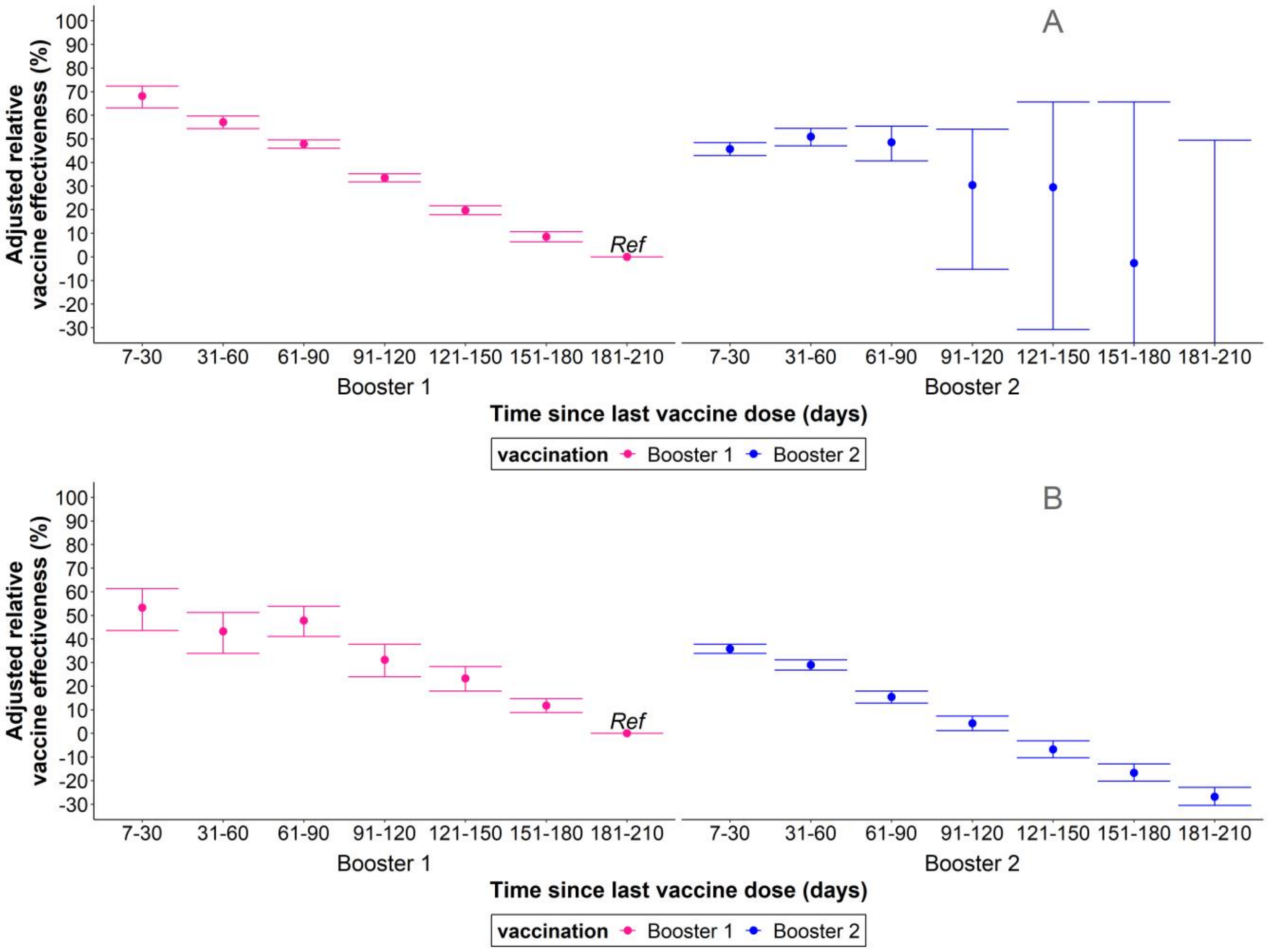


Figure S4: Adjusted relative vaccine effectiveness of the second booster of mRNA COVID-19 vaccine against **A.** Omicron BA.2 and **B.** Omicron BA.4/5 symptomatic infection among ≥60 years and older, relative to those who received the first booster dose 181-210 days ago. Error bars = 95% confidence intervals of the estimates.

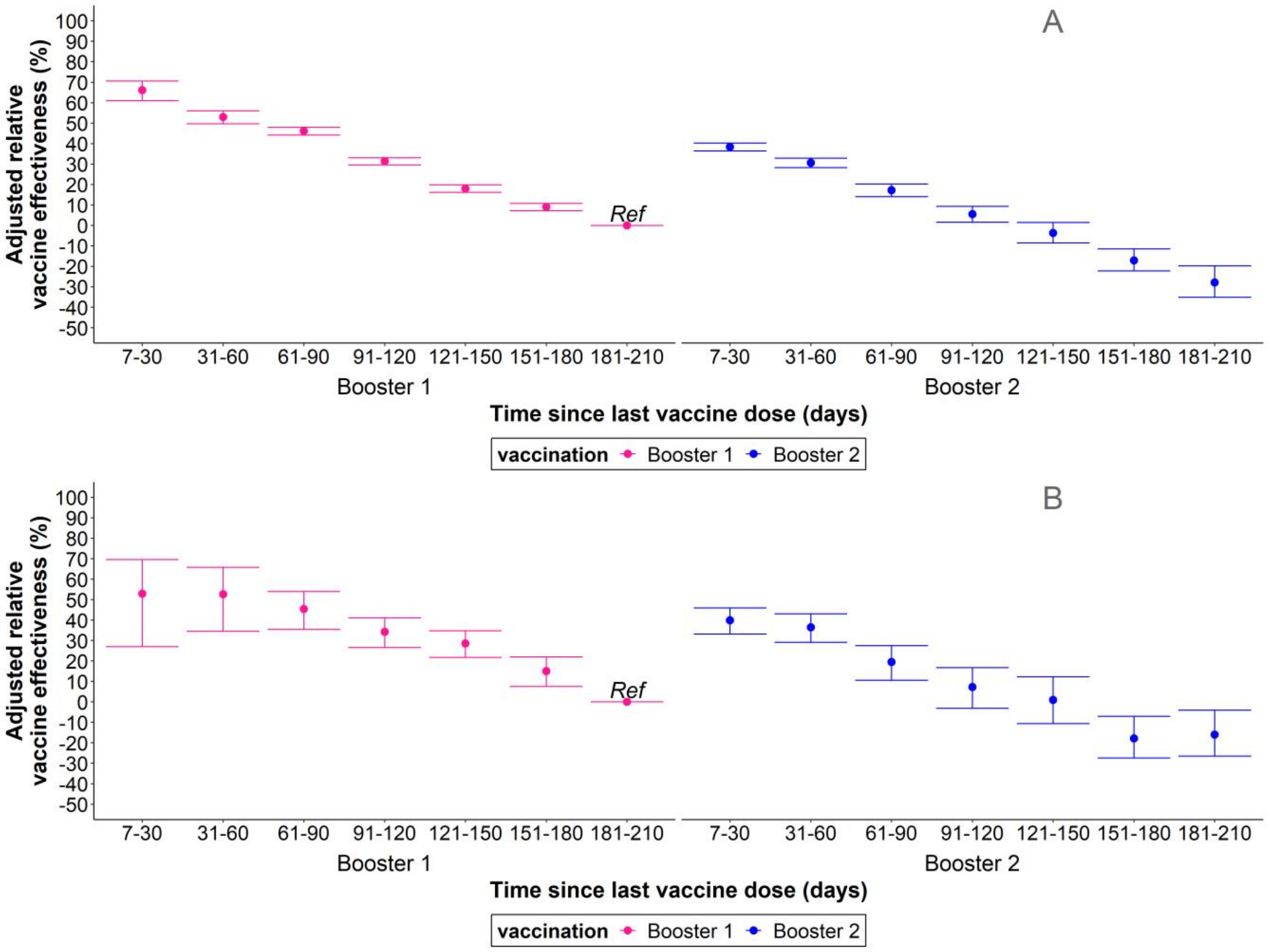


Figure S5: Adjusted relative vaccine effectiveness of the second booster of mRNA covid-19 vaccine against symptomatic Omicron BA.2 or Omicron BA.4/5 infection among **A.** 60-79 years old and **B.** ≥80 years old, relative to those who received the first booster dose 181-210 days ago. Error bars = 95% confidence intervals of the estimates.

### References

1. Cole P, MacMahon B. Attributable risk percent in case-control studies. Br J Prev Soc Med. 1971;25(4):242–4.
